# Glaucoma cases reported post-COVID-19 vaccination : A VAERS database analysis

**DOI:** 10.1101/2022.06.13.22276314

**Authors:** Rohan Bir Singh, Uday Pratap Singh Parmar, Parul Ichhpujani

**Affiliations:** Massachusetts Eye and Ear, Department of Ophthalmology, Harvard Medical School, Boston; Department of Ophthalmology, Leiden University Medical Center, Leiden, The Netherlands; Discipline of Ophthalmology and Visual Sciences, Faculty of Health and Medical Sciences, Adelaide Medical School, University of Adelaide, Australia; Glaucoma Service, Department of Ophthalmology, Government Medical College and Hospital, Chandigarh, India

## Abstract

**Objective:** To evaluate glaucoma cases reported post-COVID-19 vaccination and describe the clinical presentations in these cases.

**Design:** An analysis of the Centers for Disease Control and Prevention (CDC) Vaccine Adverse Event Reporting System (VAERS) database

**Participants:** The study includes 161 individuals who were reported for glaucoma after administration of COVID-19 vaccines [BNT162b2 (Pfizer-BioNTech), mRNA-1273 (Moderna) and Ad26.COV2.S (Janssen)] between December 2020 and April 2022.

**Main Outcome Measures:** Estimated crude reporting rate of glaucoma, clinical presentations, onset duration and associated risk factors.

**Results:** A total of 2,061,557,270 doses of COVID-19 vaccines were administered during the study timeframe. During this period, 161 glaucoma cases were reported with an estimated crude reporting rate (per million doses) of 0.09, 0.06 and 0.07 for BNT162b2, mRNA-1273 and Ad26.COV2.S, respectively. The majority of patients (n=130, 80.7%) received BNT162b2, vaccine, while 27 patients (16.8%) received mRNA-1273 and four patients (2.5%) received rAd26.COV2.S vaccines. The mean age of patients in the cohort was 60.41±17.56 years and 67.7% (n=109) were women. More than half (56.6%) were reported within the first week after vaccination. The glaucoma onset interval was significantly shorter in patients who received BNT162b2 and rAd26.COV2.S vaccines compared to mRNA-1273 (p=0.013). A higher risk of glaucoma incidence was observed in the short term in patients who received BNT162b2 vaccines compared to mRNA-1273 (p=0.05). In patients vaccinated with mRNA-1273, a significantly higher frequency of glaucoma cases was reported in older patients (p=0.047) compared to younger age groups.

**Conclusions:** The CDC-VAERS data suggest an extremely low safety concern for glaucoma on receiving BNT162b2, mRNA-1273, or Ad26.COV2.S vaccines. The onset interval of adverse events was shorter in patients who received BNT162b2 and rAd26.COV2.S vaccines compared to mRNA-1273. The glaucoma cases after mRNA-1273 vaccination were more commonly reported in older patients. These findings are subject to the limitations of passive reporting systems, under reporting and presumptive case definition, and should be considered preliminary without the medical record analysis for establishing a definitive diagnosis.

## INTRODUCTION

In response to the COVID-19 pandemic, a large-scale, global vaccination program was launched in December 2020.^1^ Since, COVID-19 vaccines were provided emergency use authorization by the US Food and Drug Administration (FDA), the Centers for Disease Control and Prevention (CDC) expanded its passive surveillance system [known as Vaccine Adverse Event Reporting System (VAERS)] to include a wide array of adverse events of special interest, including ophthalmic disorders such as glaucoma.^2^

Glaucoma is a group of ocular disorders causing characteristic optic neuropathy with corresponding visual field defects as a result of progressive degeneration of retinal ganglion cells in the optic disc and loss of their axons in the optic nerve.^3^ Raised intraocular pressure (IOP) has been identified as a major risk factor.These pathological changes lead to progressive peripheral visual field defects and may eventually result in blindness. Despite the thorough clinical insights into primary as well as secondary glaucoma, the pathological mechanisms at the cellular and sub-cellular level are poorly understood and the factors significantly contributing to disease progression are yet to be delineated.^4^ In 2020, an estimated 76.02 million people were reported to have glaucoma worldwide.^5^ The COVID-19 pandemic has had an enormous impact on glaucoma care globally, impeding diagnosis, patient follow-up and surgeries.^6^ Although, there are reports highlighting ocular adverse events associated with COVID-19 vaccination, the literature is sparse associating COVID-19 vaccination with glaucoma. Moreover, the COVID-19 vaccine related information portals categorically report the absence of evidence linking vaccination with glaucoma.

We used the VAERS database to evaluate the reports of glaucoma cases for three FDA-approved COVID-19 vaccines: BNT162b2 (Pfizer BioNtech), mRNA-1273 (Moderna), and Ad26.COV2.S (Janssen). We determine the crude reporting rate of glaucoma in vaccine recipients since the initiation of vaccination program and compare these rates for the three vaccines. We report the clinical characteristics in patients diagnosed with glaucoma and assessed the association between age, sex, and duration of onset (post-vaccination) in the patients who received either of the three vaccines.

## METHODS

This study includes publicly available, de-identified, anonymous data and does not require approval by the institutional review board. VAERS is a passive surveillance system that functions as an early warning system for potential vaccine adverse events.^2^ The VAERS data are available through Wide-ranging Online Data for Epidemiologic Research (WONDER), a database developed and operated by the Centers for Disease Control and Prevention, an agency of the United States federal government. The VAERS database compiles reports of all post-vaccination adverse events from patients, parents (for minor patients), clinicians, vaccine manufacturers, and regulatory bodies globally. The database includes a detailed report on the post-vaccination adverse events experienced by the patient after vaccination. The data recorded in VAERS are verified by third-party professional coders who assign appropriate medical terminology (on basis of Medical Dictionary for Regulatory Activities preferred terms) from the data in the submitted reports.^7^ CDC WONDER allows access to the information freely, and use, copy, distributing or publishing this information without additional or explicit permission.^8^ The recorded data included twenty-two labelled and eight unspecified countries. The glaucoma related adverse effects data from the United States were reported from 26 states.

The VAERS data included in this study were accessed via CDC WONDER on April 30^th^, 2022.^9^ The data query included all COVID-19 vaccine adverse events for all vaccine types administered to patients of all ages and genders for glaucoma (unspecified type), angle closure glaucoma, open angle glaucoma, uveitic glaucoma, uveitis-glaucoma-hyphaema syndrome. The results were grouped by symptoms, age, sex, state/territory and onset interval. The additional measures included in the results were adverse event description, lab data, current illness, adverse events after prior vaccination, medications at time of vaccination and history/allergies. The data included in the analysis were clinical presentation, date of vaccination and adverse event onset, ocular and systemic history along with prescribed drugs and surgeries performed in the patients prior to presentation. Some of the patient reports also included the interventions post-glaucoma diagnosis.

The unspecified glaucoma data were stratified broadly into open angle glaucoma (OAG) and angle closure glaucoma (ACG), as per the reported clinical presentation by a glaucoma specialist (PI). The patients presenting with elevated IOP and general ocular symptoms such as eye pain, without any associated ocular morbidity were considered as open angle glaucoma. The patients who were categorically reported with vision loss/blindness associated with colored haloes and redness, were classified as angle closure glaucoma. Although there may be some discrepancies in this classification as the gonioscopy findings were not mentioned. The crude reporting rates were estimated by using the number of glaucoma reports (by vaccine type) per million COVID-19 vaccine doses.

## STATISTICAL ANALYSIS

The statistical analysis was performed using R Studio (R Foundation for Statistical Computing, Vienna, Austria). The crude reporting rates were estimated by using the number of glaucoma reports (by vaccine type) per million COVID-19 vaccine doses. We performed a descriptive analysis of the social demographic characteristics and vaccination data. We assessed the association between onset interval of glaucoma and vaccine type, age, sex, and dosage using the ANOVA test. The t-test was used to evaluate the association between glaucoma diagnosis and prior history of COVID-19. A reverse Kaplan-Meier risk analysis was performed for the two vaccines - BNT162b2 and mRNA-1273.The Ad26.COV2.S vaccine was excluded from this analysis due to few reports. Since the primary outcome measure (i.e. glaucoma diagnosis) was categorical, the analysis was performed to investigate risk factors associated with it using Pearson’s Chi square test of association. The missing values in the data were indicated and Na.rm code to account for it during the analysis

## RESULTS

A total of 2,061,557,270 COVID-19 vaccine doses were administered during the study period and 80.7% were BNT162b2, 16.8% were mRNA-1273 and 2.5% were Ad26.COV2.S.^1^ During this period, 1,250,310 (0.06% of all doses) adverse events following COVID-19 vaccinations were recorded in CDC VAERS, including 166 reports of glaucoma.^2^ In our analysis, 161 reports were included, four reports were duplicate, and one report did not include any information except the type of vaccine administered. The cases were reported by drug regulatory agencies (n=99, 61.5%), physicians (n=18, 11.2%), directly by patients (n=26, 16.1%) and vaccine manufacturers (n=16, 9.9%). The estimated crude reporting rates (per million doses) for BNT162b2, mRNA-1273 and Ad26.COV2.S were of 0.09, 0.06 and 0.07, respectively All the cases in the cohort were classified as “medically significant” adverse event by the CDC VAERS. The average age of the patients included in the study was 60.41±17.56 years and 67.7% (n=109) were female. **(Table 1)** The cases were reported from the United States (48, 29.69%), Europe (86, 51.8%), and Asia (18, 10.8%), respectively. **(Supplementary Table 1)** The state-by-state crude reporting rate for the three vaccines administered in the United States is reported in **Table 2**.

**Table 1:**
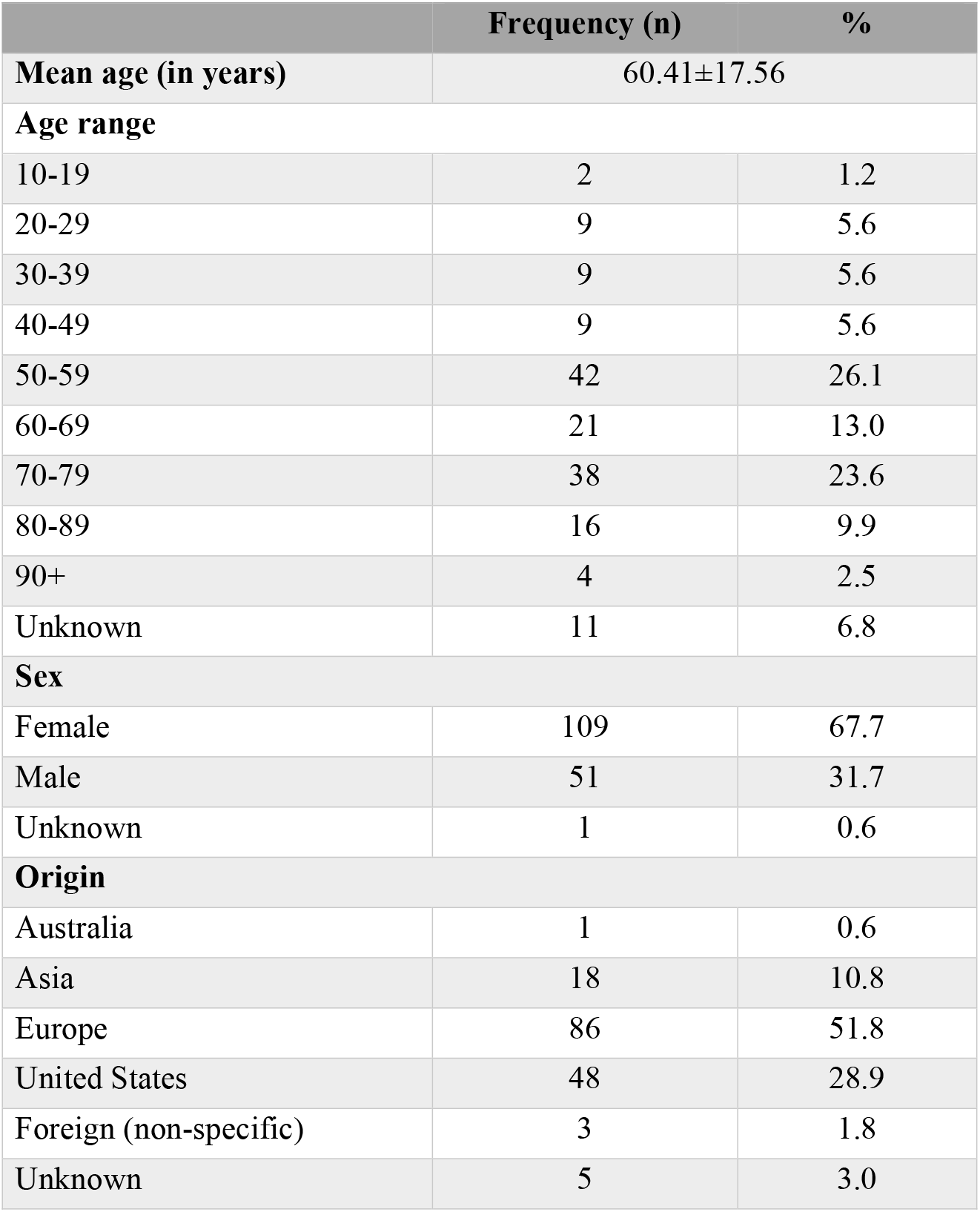
The demographics with patients who were reported with glaucoma post-COVID-19 vaccination

**Table 2:**
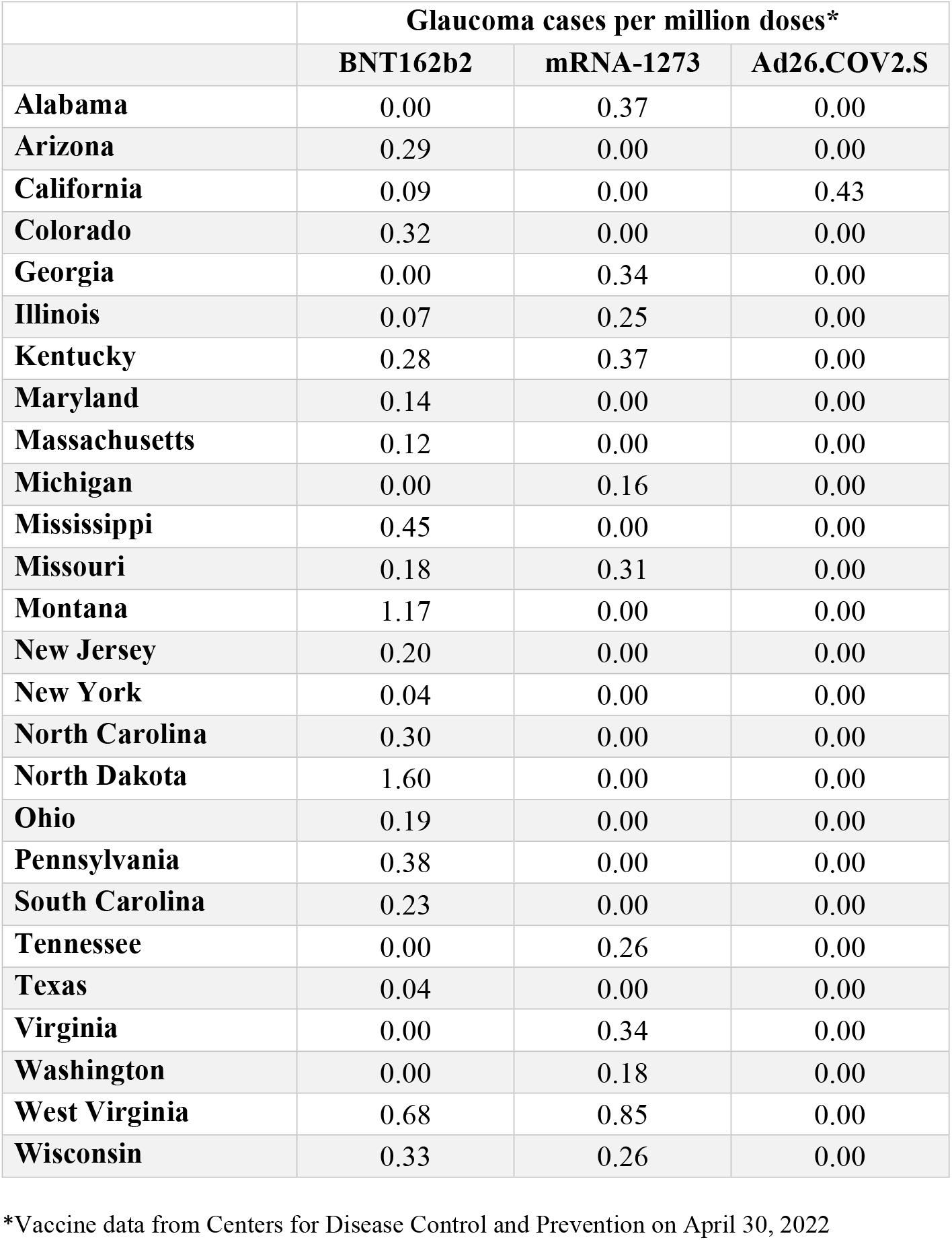
State-wise crude cases of glaucoma per million doses of COVID-19 vaccinations

|  | **Glaucoma cases per million doses\*** |  |  |
| --- | --- | --- | --- |
|  | **BNT162b2** | **mRNA-1273** | **Ad26.COV2.S** |
| **Alabama** | 0.00 | 0.37 | 0.00 |
| **Arizona** | 0.29 | 0.00 | 0.00 |
| **California** | 0.09 | 0.00 | 0.43 |
| **Colorado** | 0.32 | 0.00 | 0.00 |
| **Georgia** | 0.00 | 0.34 | 0.00 |
| **Illinois** | 0.07 | 0.25 | 0.00 |
| **Kentucky** | 0.28 | 0.37 | 0.00 |
| **Maryland** | 0.14 | 0.00 | 0.00 |
| **Massachusetts** | 0.12 | 0.00 | 0.00 |
| **Michigan** | 0.00 | 0.16 | 0.00 |
| **Mississippi** | 0.45 | 0.00 | 0.00 |
| **Missouri** | 0.18 | 0.31 | 0.00 |
| **Montana** | 1.17 | 0.00 | 0.00 |
| **New Jersey** | 0.20 | 0.00 | 0.00 |
| **New York** | 0.04 | 0.00 | 0.00 |
| **North Carolina** | 0.30 | 0.00 | 0.00 |
| **North Dakota** | 1.60 | 0.00 | 0.00 |
| **Ohio** | 0.19 | 0.00 | 0.00 |
| **Pennsylvania** | 0.38 | 0.00 | 0.00 |
| **South Carolina** | 0.23 | 0.00 | 0.00 |
| **Tennessee** | 0.00 | 0.26 | 0.00 |
| **Texas** | 0.04 | 0.00 | 0.00 |
| **Virginia** | 0.00 | 0.34 | 0.00 |
| **Washington** | 0.00 | 0.18 | 0.00 |
| **West Virginia** | 0.68 | 0.85 | 0.00 |
| **Wisconsin** | 0.33 | 0.26 | 0.00 |

\*Vaccine data from Centers for Disease Control and Prevention on April 30, 2022

In the study cohort, majority of the patients (n=130, 80.7%) were administered BNT162b2, vaccine, while 27 patients (16.8%) received mRNA-1273 and four patients (2.5%) received rAd26.COV2.S vaccines. **(Table 3)** Most of the cases (n=91, 56.5%) were reported within first week, including 18% (n=29) of the cases on the day of vaccination. **(Figure 1)** Seventy-seven cases (47.8%) were reported after the first dose, 59 cases (36.6%) were reported after the second dose and 13 cases (8.1%) were reported after the third dose of the vaccine. **(Figure 2)** Only five patients (3.1%) reported a prior history of COVID-19. On stratifying the patients based on clinical descriptions, we found that most of the cases (n=105, 65.2%) had OAG. The patients presented with ocular pain (n=60, 37.3%), reduced/blurry vision (47, 29.2%), and complete vision loss/blindness (n=35, 21.7%). An elevated IOP was reported in 48 (29.8%) cases. The other ocular signs included flashes (n=6, 3.7%), floaters (n=5, 3.1%), and photophobia (n=5, 3.1%). Notably, 28 patients (17.3%) had a prior history of glaucoma and had controlled IOP at the time of vaccination. Seven patients (4.3%) had a history of uveitis. The patients also presented with severe headache (n=48, 29.8%), general body pain (n=17, 10.6%), and high blood pressure (n=8, 5.0%). The patients had a prior history of cardiovascular diseases (n=33, 20.5%), hypertension (n=13, 8.1%). The ocular and systemic presentation and history of the patients included in the study is summarized in **table 4**.

**Table 3:**
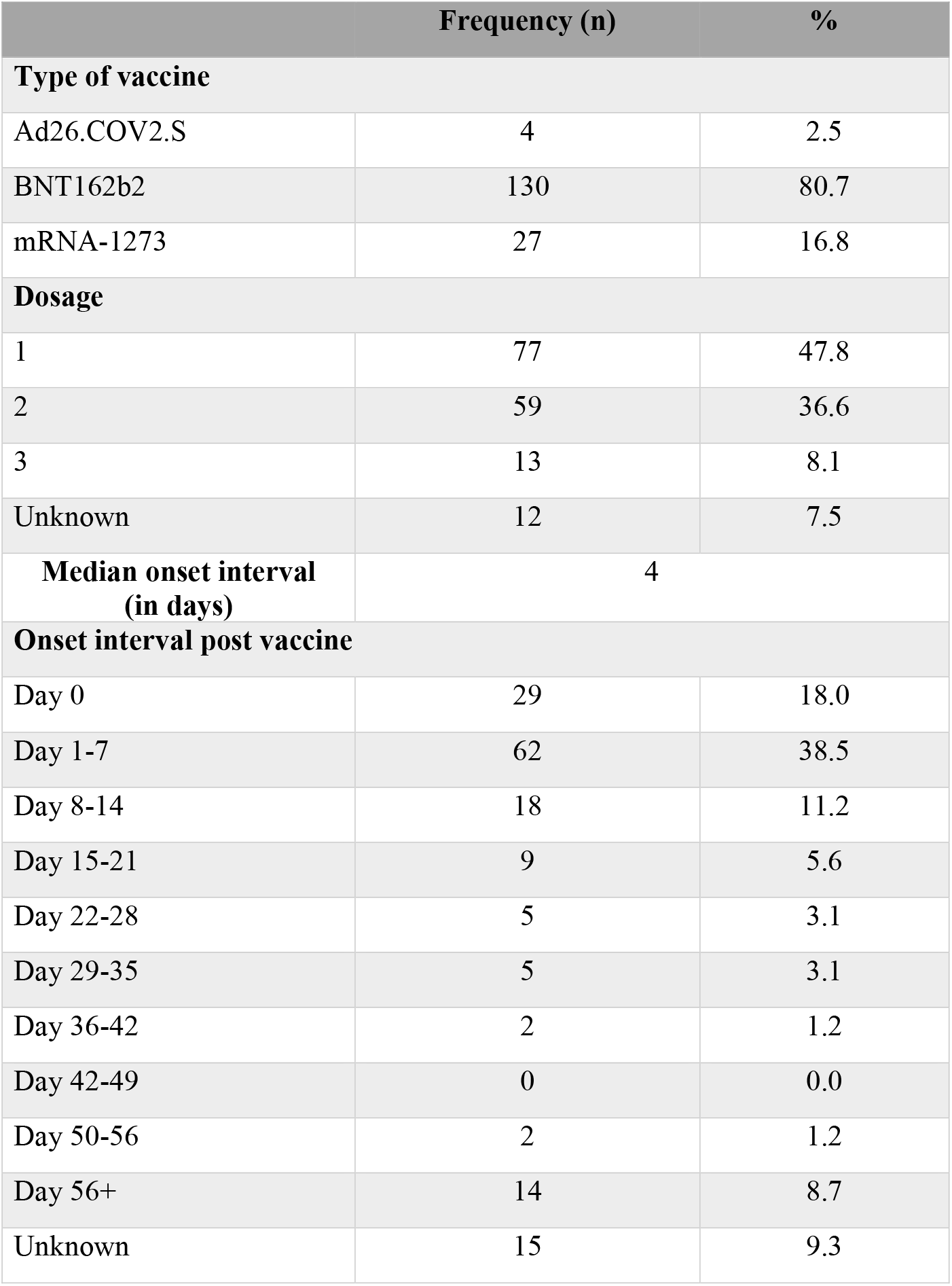
The vaccination data of the patients who were reported with glaucoma post-vaccination

**Table 4:** The ocular and systemic history and presentation in patients who were reported with glaucoma post-COVID-19 vaccinations

|  | Frequency | % |
| --- | --- | --- |
| **History of COVID-19** | 5 | 3.1 |
| **Glaucoma diagnosis** |  |  |
| Open angle glaucoma | 105 | 65.2 |
| Angle closure glaucoma | 45 | 27.6 |
| Unknown | 11 | 6.8 |
| **Ocular Presentation** |  |  |
| Flashes | 6 | 3.7 |
| Floaters | 5 | 3.1 |
| High IOP | 48 | 29.8 |
| Ocular pain | 60 | 37.3 |
| Photophobia | 5 | 3.1 |
| Reduced/Blurry vision | 47 | 29.2 |
| Vision loss/Blindness | 35 | 21.7 |
| **Ocular history** |  |  |
| Conjunctivitis | 4 | 2.5 |
| Glaucoma (controlled) | 28 | 17.3 |
| Hemorrhage | 3 | 1.9 |
| Herpes Zoster Ophthalmicus | 5 | 3.1 |
| Keratitis | 2 | 1.2 |
| Ocular ischemic syndrome | 1 | 0.6 |
| Optic ischemic neuropathy | 1 | 0.6 |
| Retinal artery occlusion | 1 | 0.6 |
| Retinal vein occlusion | 6 | 3.7 |
| Uveitis | 7 | 4.3 |
| Vitreous detachment | 2 | 1.2 |
| **Systemic Presentation** |  |  |
| Headache | 48 | 29.8 |
| Pain | 17 | 10.6 |
| Nausea | 12 | 7.5 |
| Palpitations | 4 | 2.5 |
| High Blood Pressure | 8 | 5.0 |
| Vaccine site induration/rash | 6 | 3.7 |
| **Systemic history** |  |  |
| Allergies | 9 | 5.6 |
| Cardiovascular Disorders (MI, CAD, Afib, Tachyarrhythmia, Heart failure) | 33 | 20.5 |
| Cerebrovascular Disorders | 3 | 1.9 |
| Diabetes | 9 | 5.6 |
| Hypercholesterolemia/Dyslipidemia | 7 | 4.3 |
| Hypertension | 13 | 8.1 |
| Hypothyroidism | 11 | 6.8 |
| Pulmonary Disorders (COPD,<br>Embolism) | 4 | 2.5 |
| Renal Disorders | 5 | 3.1 |

**Table 5:** Analysis to assess the factors associated with onset interval of glaucoma post-COVID-19 vaccination

|  | Percentage (n) | Mean onset interval<br>(in days) | p-value |
| --- | --- | --- | --- |
| Vaccine* |  |  |  |
| BNT162b2 | 80.7% (130/161) | 14.7 ± 27.58 | 0.013* |
| mRNA-1273 | 16.7% (27/161) | 37.07 ± 66.11 |  |
| Ad26.COV2.S | 2.5% (4/161) | 5.5 ± 6.4 |  |
| Sex* |  |  |  |
| Female | 67.7% (109/161) | 15.17 ± 31.2 | 0.196 |
| Male | 31.7% (51/161) | 24.6 ± 47.5 |  |
| Age* |  |  |  |
| 10-19 | 1.86% (3/161) | 32 ± 53.69 | 0.565 |
| 20-29 | 5.6% (9/161) | 21.88 ± 53.24 |  |
| 30-39 | 6.8% (11/161) | 29.81 ± 77.93 |  |
| 40-49 | 8% (13/161) | 12.46 ± 17.83 |  |
| 50-59 | 32.3% (52/161) | 14.90 ± 26.29 |  |
| 60-69 | 13% (21/161) | 27.04 ± 39.48 |  |
| 70-79 | 21.7% (35/161) | 15.57 ± 30.93 |  |
| 80-89 | 8.7% (14/161) | 20.71 ± 45.39 |  |
| 90+ | 1.86% (3/161) | 12 ± 18.19 |  |
| Dosage* |  |  |  |
| 1 | 47.8% (77/161) | 15.34 ± 37.35 | 0.268 |
| 2 | 36.6% (59/161) | 23.64 ± 43.03 |  |
| 3 | 8.07% (13/161) | 17.61 ± 22.86 |  |
| Unknown | 7.45% (12/161) |  |  |
| History of COVID-19** |  |  |  |
| Yes | 3.1% (5/161) | 54.2 ± 72.8 | 0.08 |
| No | 96.27% (155/161) | 17.18 ± 35.63 |  |
| Unknown | 0.62% (1/161) |  |  |

\* One-way ANOVA test, \*\*T-test

**Table 6:**
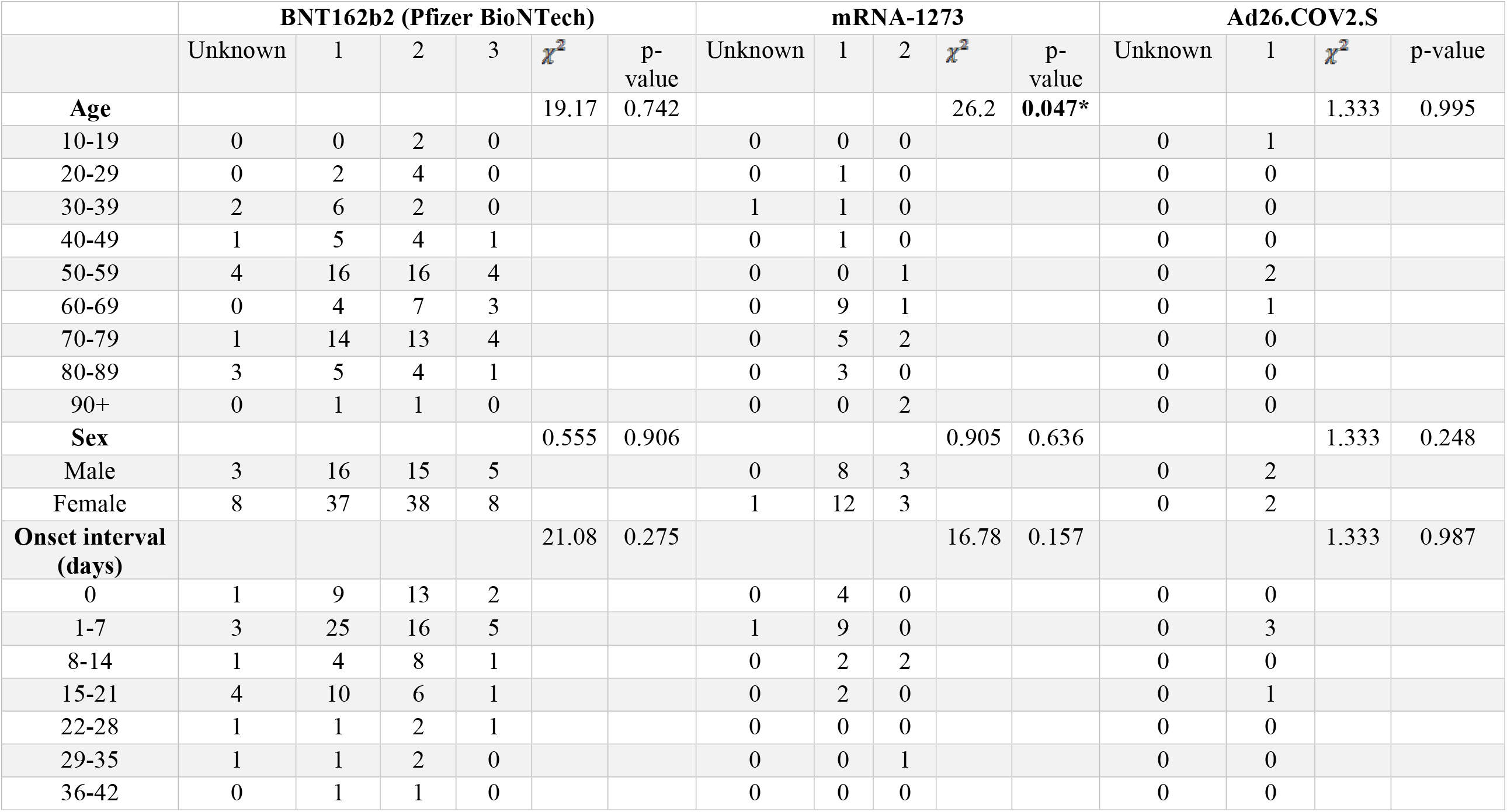
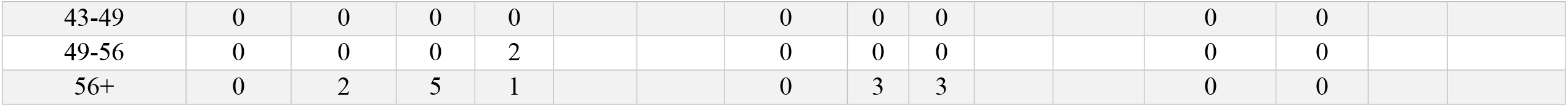
Association analysis of age, sex, onset interval with glaucoma post-COVID-19 vaccination

|  | BNT162b2 (Pfizer BioNTech) |  |  |  |  |  | mRNA-1273 |  |  |  |  | Ad26.COV2.S |  |  |  |
| --- | --- | --- | --- | --- | --- | --- | --- | --- | --- | --- | --- | --- | --- | --- | --- |
| | Unknown | 1 | 2 | 3 | $\chi^2$ | p-value | Unknown | 1 | 2 | $\chi^2$ | p-value | Unknown | 1 | $\chi^2$ | p-value |
| Age |  |  |  |  | 19.17 | 0.742 |  |  |  | 26.2 | 0.047* |  |  | 1.333 | 0.995 |
| 10-19 | 0 | 0 | 2 | 0 |  |  | 0 | 0 | 0 |  |  | 0 | 1 |  |  |
| 20-29 | 0 | 2 | 4 | 0 |  |  | 0 | 1 | 0 |  |  | 0 | 0 |  |  |
| 30-39 | 2 | 6 | 2 | 0 |  |  | 1 | 1 | 0 |  |  | 0 | 0 |  |  |
| 40-49 | 1 | 5 | 4 | 1 |  |  | 0 | 1 | 0 |  |  | 0 | 0 |  |  |
| 50-59 | 4 | 16 | 16 | 4 |  |  | 0 | 0 | 1 |  |  | 0 | 2 |  |  |
| 60-69 | 0 | 4 | 7 | 3 |  |  | 0 | 9 | 1 |  |  | 0 | 1 |  |  |
| 70-79 | 1 | 14 | 13 | 4 |  |  | 0 | 5 | 2 |  |  | 0 | 0 |  |  |
| 80-89 | 3 | 5 | 4 | 1 |  |  | 0 | 3 | 0 |  |  | 0 | 0 |  |  |
| 90+ | 0 | 1 | 1 | 0 |  |  | 0 | 0 | 2 |  |  | 0 | 0 |  |  |
| Sex |  |  |  |  | 0.555 | 0.906 |  |  |  | 0.905 | 0.636 |  |  | 1.333 | 0.248 |
| Male | 3 | 16 | 15 | 5 |  |  | 0 | 8 | 3 |  |  | 0 | 2 |  |  |
| Female | 8 | 37 | 38 | 8 |  |  | 1 | 12 | 3 |  |  | 0 | 2 |  |  |
| Onset interval (days) |  |  |  |  | 21.08 | 0.275 |  |  |  | 16.78 | 0.157 |  |  | 1.333 | 0.987 |
| 0 | 1 | 9 | 13 | 2 |  |  | 0 | 4 | 0 |  |  | 0 | 0 |  |  |
| 1-7 | 3 | 25 | 16 | 5 |  |  | 1 | 9 | 0 |  |  | 0 | 3 |  |  |
| 8-14 | 1 | 4 | 8 | 1 |  |  | 0 | 2 | 2 |  |  | 0 | 0 |  |  |
| 15-21 | 4 | 10 | 6 | 1 |  |  | 0 | 2 | 0 |  |  | 0 | 1 |  |  |
| 22-28 | 1 | 1 | 2 | 1 |  |  | 0 | 0 | 0 |  |  | 0 | 0 |  |  |
| 29-35 | 1 | 1 | 2 | 0 |  |  | 0 | 0 | 1 |  |  | 0 | 0 |  |  |
| 36-42 | 0 | 1 | 1 | 0 |  |  | 0 | 0 | 0 |  |  | 0 | 0 |  |  |
| 43-49 | 0 | 0 | 0 | 0 |  |  | 0 | 0 | 0 |  |  | 0 | 0 |  |  |
| 49-56 | 0 | 0 | 0 | 2 |  |  | 0 | 0 | 0 |  |  | 0 | 0 |  |  |
| 56+ | 0 | 2 | 5 | 1 |  |  | 0 | 3 | 3 |  |  | 0 | 0 |  |  |

**Table 7:** Therapeutic interventions reported in patients diagnosed with glaucoma post-COVID-19 vaccination

|  | Frequency | % |
| --- | --- | --- |
| Brimonidine | 5 | 3.1 |
| Dorzolamide | 6 | 3.7 |
| Eye drops (not specified) | 18 | 11.2 |
| Laser iridotomy | 18 | 11.2 |
| Mannitol | 2 | 1.2 |
| Shunt/Valve placement | 4 | 2.5 |
| Surgery (not specified) | 7 | 4.3 |
| Timolol | 11 | 6.8 |
| Trabeculectomy | 2 | 1.2 |
| Travoprost/Latanoprost | 2 | 1.2 |
| No/unknown intervention | 86 | 53.4 |

**Figure 1:**
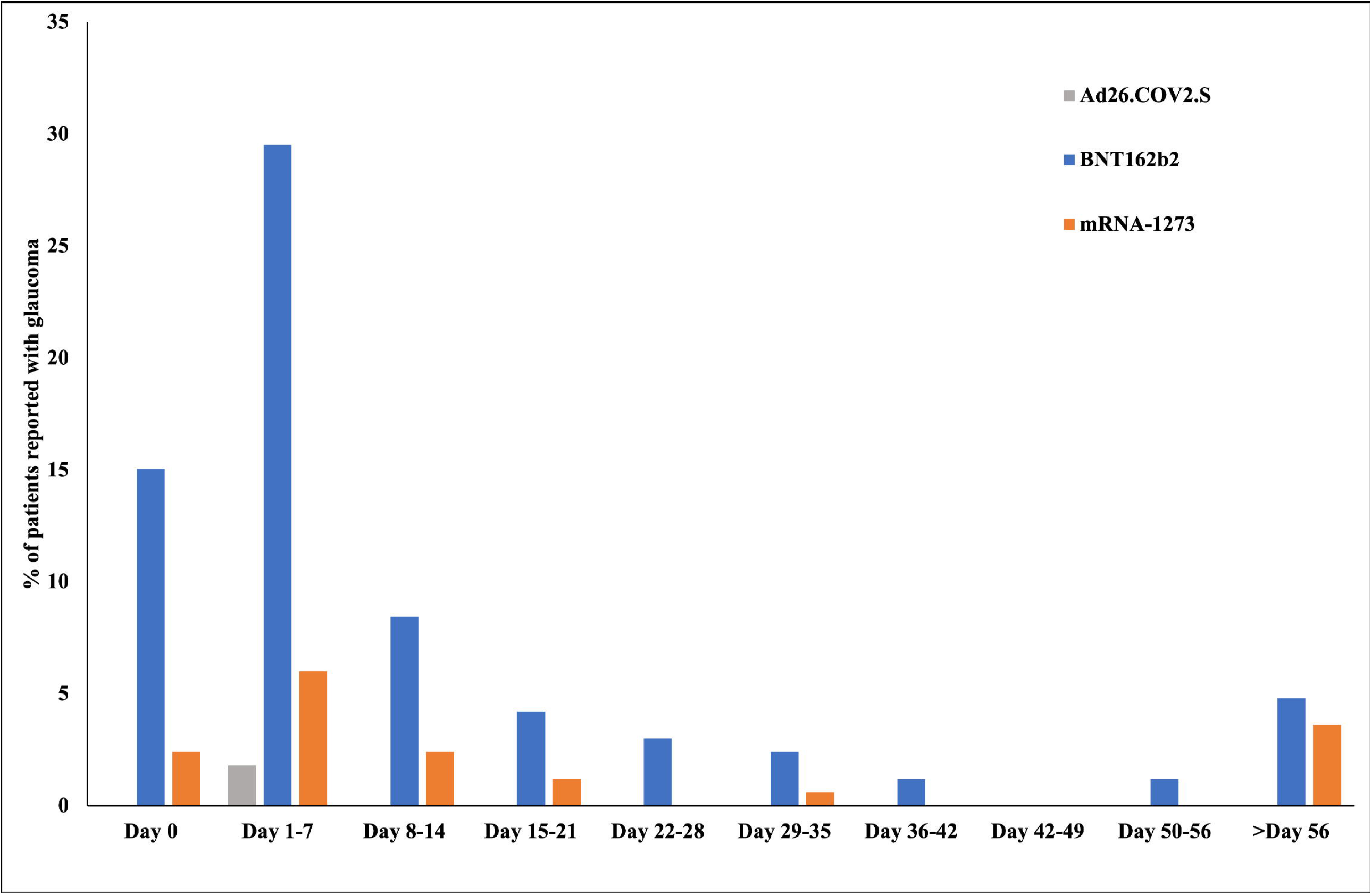
The cases of glaucoma post-vaccination with BNT162b2, mRNA-1273 and Ad26.COV2.S on day 0 (i.e., day of vaccination) and in subsequent weeks

**Figure 2:**
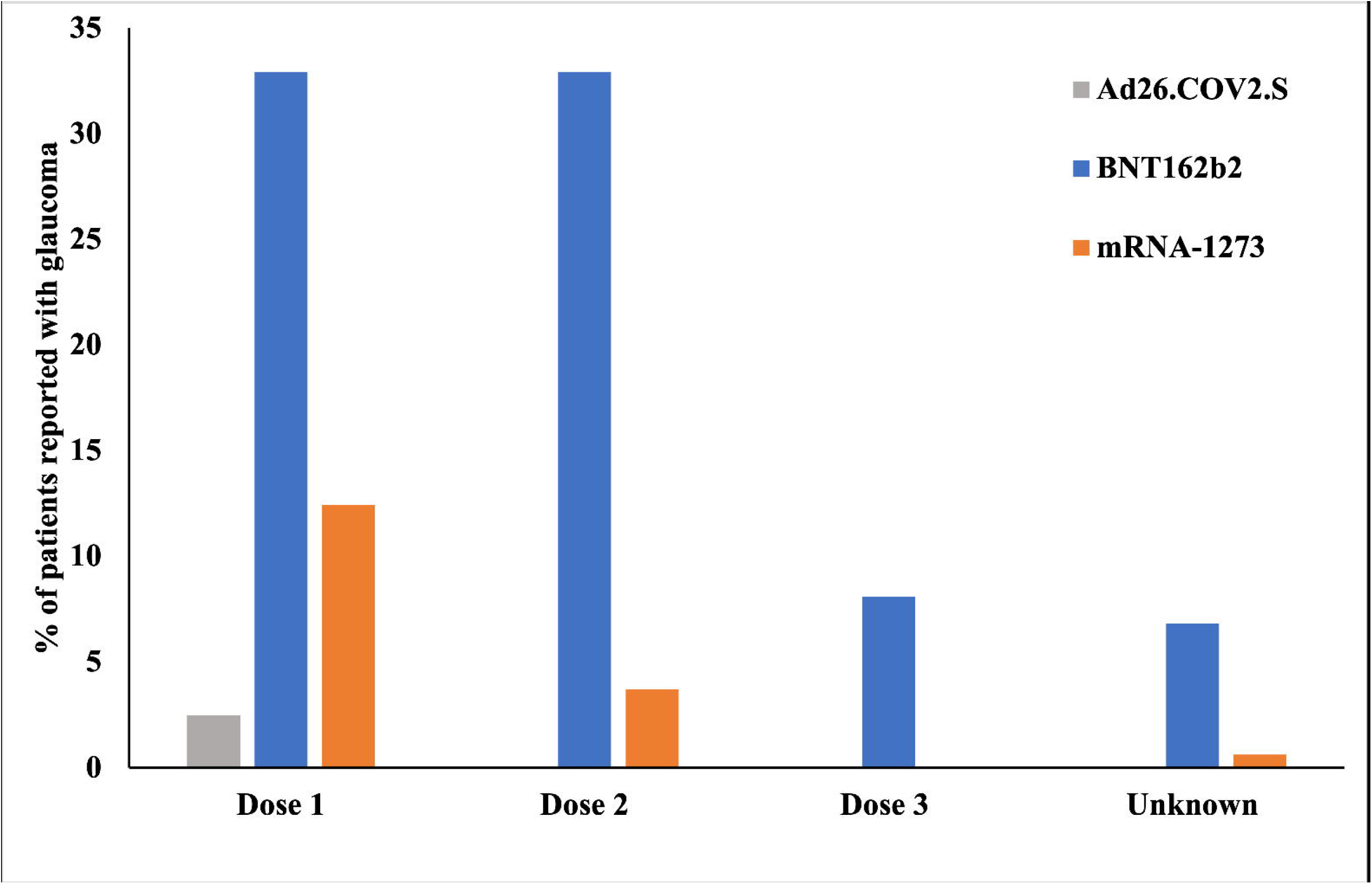
The cases of glaucoma post vaccination with protocol doses (dose 1 and 2 for BNT162b2, mRNA-1273 and dose 1 for Ad26.COV2.S) and boosters.

**Figure 3:**
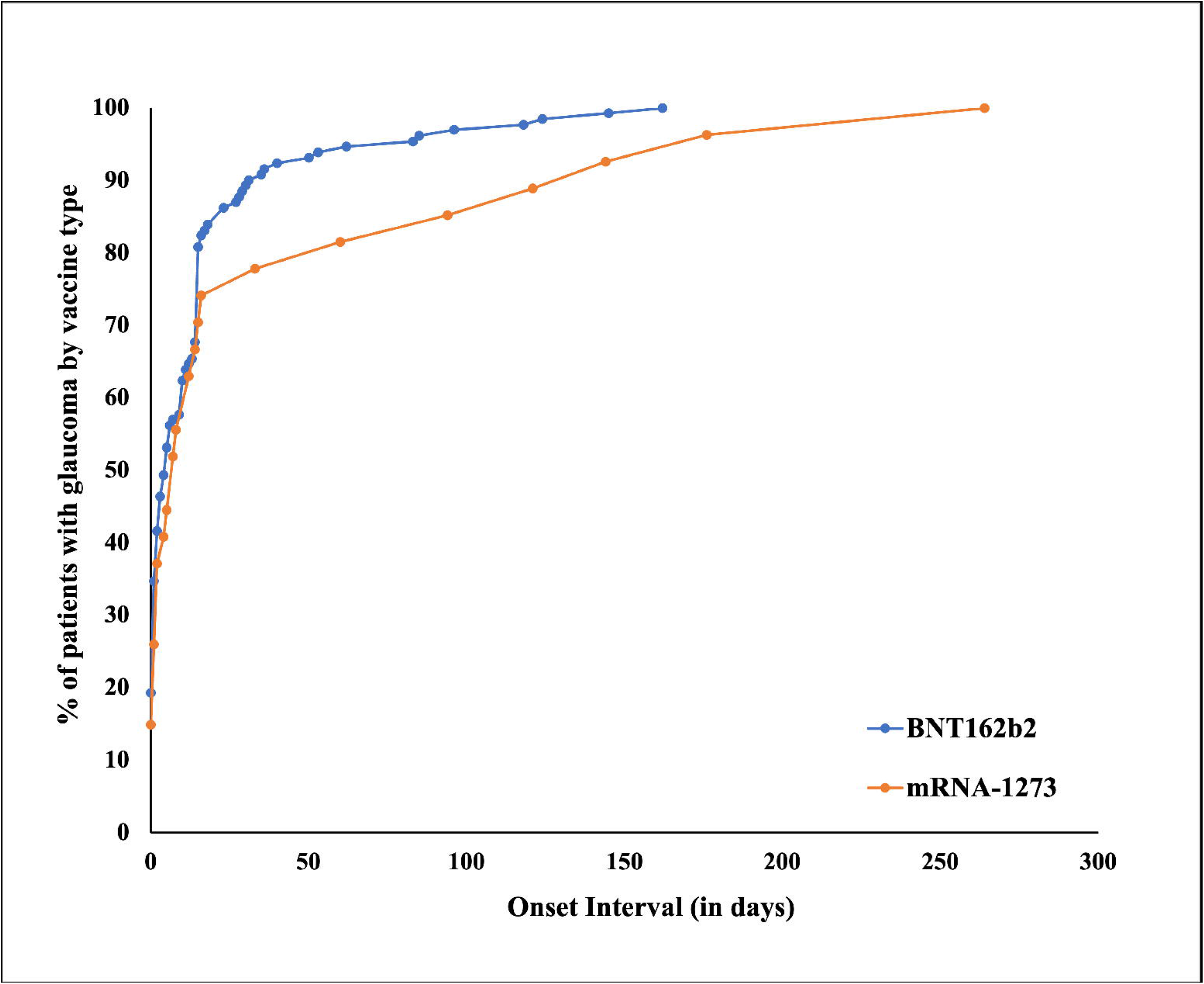
Reverse Kaplan-Meier risk analysis for glaucoma cases report after administration of BNT162b2 and mRNA-1273 vaccines.

The statistical evaluation revealed an association between vaccine type and duration of glaucoma presentation post-vaccination. The time of symptom onset in patients who received BNT162b2 (14.7 ± 27.58 days) and Ad26.COV2.S (5.5 ± 6.4 days) vaccines was significantly shorter compared to mRNA-1273 (37.07 ± 66.11 days, p=0.013). These findings were confirmed by reverse Kaplan-Meier risk analysis, which showed a significant difference in the onset duration between BNT162b2 and mRNA-1273 (p=0.05). The analysis showed that there was no significant association between onset interval and sex (p=0.196), age (0.565), and history of COVID-19 (p=0.08). The Pearson’s Chi-square analysis showed that the frequency of glaucoma cases in patients who received mRNA-1273 vaccine was significantly higher (p=0.047) in the older age groups (6^th^-7^th^ decade). A similar association was not observed in patients who were vaccinated with BNT162b2 and Ad26.COV2.S vaccines. Additionally, we did not observe any significant association between the vaccine doses, sex and onset interval.

The patients were prescribed topical eye drops (n=44) and surgical interventions (n=29) were performed in requisite cases. Laser iridotomy and shunt/valve placement surgeries were performed in 18 (11.2%) and 4 (2.5%) patients, respectively. In this cohort, none of the patients had a pre-existing iridotomy. Trabeculectomy was performed in two (1.2%) patients. The type of surgical intervention was not specified in seven (4.3%) patients. The patients were prescribed brimonidine (n=5, 3.1%), dorzolamide (n=6, 3.7%), timolol (n=11, 6.8%), or travoprost/latanoprost (n=2, 1.2%). The prescribed eye drops were not specified in 18 (11.2%) patients. The interventions were not known for more than half (n=86;53.4%) patients.

## DISCUSSION

Since the initiation of one of the largest global vaccination programs in December 2020, several case reports have highlighted the ocular adverse events associated with the COVID-19 vaccines. The Bradford-Hill criteria include nine aspects to consider when inferring causality between events: strength of the association, consistency, specificity, temporality, biological gradient, plausibility, coherence, experiment, and analogy. The current study ensures temporality, as there was a short interval of time following vaccination and the onset of the signs for glaucoma.^10^

The ocular adverse events associated with the three vaccines approved in the United States include Bell’s and abducens nerve palsy, acute macular neuroretinopathy, central serous retinopathy, Grave’s disease, Vogt-Koyanagi-Harada disease, retinal and ophthalmic vein thrombosis and corneal graft rejection.^11–19^ Similar ocular disorders have also been observed in patients who are infected by COVID-19 virus.^20–22^ However, only two cases of secondary angle closure glaucoma post-vaccination have been reported in the literature. Behera and colleagues reported the case of a 60-year-old male with hemophilia who developed painful and sudden vision loss in his eye a day after receiving ChAdOx1 nCoV-19 (Oxford AstraZeneca) vaccine due to acute angle-closure glaucoma secondary to a massive suprachoroidal hemorrhage.^23^ In the other case, a 49-year-old male presented with progressive vision loss one day after administration of the second dose of BNT162b2 vaccine. The patient had a massive intraocular hemorrhage and was diagnosed with secondary angle closure glaucoma, bullous retinal detachment, and massive intraocular hemorrhage.^24^ The patient’s presentation was attributed to the necrosis of a melanocytic lesion at the posterior edge of the ciliary body and choroid.^24^ In another case report, Santovito and Pinna reported reduced vision, severe headache and photophobia in a patient following BNT162b2 vaccine. The patient had no prior ocular history, and the authors could not establish a definitive diagnosis in the patient.^13^

The analysis of the VAERS data suggest an extremely low safety concern for glaucoma following receipt of the BNT162b2, mRNA-1273, or Ad26.COV2.S vaccines. The estimated crude reporting rates in this study are comparable to the report by Wang and colleagues, who evaluated the data from the Australia Therapeutic Goods Administration Database of Adverse Event Notifications, the Canada Vigilance Adverse Reaction Database, the European Union Medicines Agency (EudraVigilance) System, and the United Kingdom Medicines and Healthcare Products Regulatory Agency between December 2020 and August 2021.^25^

In this study cohort, a substantial proportion of the patients were women and between 50-70 years. The data analysis of this cohort shows that the patients typically presented with signs of glaucoma in the first 24-48 hours post-vaccination and the incidence was higher after the first dose. Most of the patients in this cohort did not have a prior history of glaucoma, therefore it is imperative that patients at the risk of developing glaucoma should remain vigilant post-vaccination. The onset interval of the disease was significantly shorter in patients vaccinated with BNT162b2 and Ad26.COV2.S vaccine compared to the mRNA-1273 vaccine. The patients who were diagnosed with glaucoma after mRNA-1273 vaccination were primarily between 60-80 years old. Although, uveitis and other ocular inflammatory disorders are considered the more common ocular adverse events associated post-vaccination, in this cohort only 4% patients were diagnosed with it. The pathogenesis of post-vaccination secondary glaucoma can be explained by the underlying disorders. However, there are no insights into the pathophysiologic mechanisms that can potentially cause post-vaccination open angle glaucoma, reported in many of the patients included in this study.

## LIMITATIONS

This study reporting the glaucoma cases post-COVID-19 vaccines has several limitations. VAERS is a passive surveillance system with reporting by pharmaceutical companies, physicians, drug regulators and patients from all over the world. Despite the mandatory requirement to report vaccine associated adverse events, underreporting and delayed reporting are common. In some cases, the submitted reports are incomplete, and lack a uniformity in data reporting. Several reports have missing data points such as ethnicity, which are considered important risk factors associated with glaucoma.

The absence of an unvaccinated control group impedes relative risk calculation. The pharmacovigilance associated with COVID-19 vaccines is limited to the European Union, United States, Australia, Canada, and a few Asian countries and hence the reports are not recorded from many developing countries, including India where over one billion doses of vaccine have been administered. Moreover, the data are absent for several other approved vaccines such as ChAdOx1 nCoV-19, ZyCoV-D, Sputnik, Covidecia, Sputnik, Sinopharm, Abdala, Soberna, Zifivax, and Novavax. Many Bradford-Hill criteria are not met as most of the data cannot be used to infer causality as it does not allow to establish strength of the association, biological gradient, coherence, or specificity.

The reports submitted by drug regulators, pharmaceutical companies, and physicians (∼80%) can be relied on for clinical diagnosis for glaucoma, however in cases where patients have self-reported are presumptively considered to be glaucoma. The data in the literature suggests glaucoma underreporting, therefore it can be assumed that cases were grossly underreported. The lack of clinical (specifically vision and gonioscopy) data severely impedes the use of these data to ability of the researchers to convincingly report the adverse effects in these reports. Additionally, there are no details of optic nerve head cupping or visual field defects on perimetry, thus what is labeled as glaucoma can also be secondary ocular hypertension, as the access to detailed clinical history is not logistically possible and is limited by the Health Insurance Portability and Accountability Act (HIPPA) of 1996.^26^

## Data Availability

All data produced in the present study are available upon reasonable request to the authors

## TABLE CAPTIONS

**Supplementary Table 1:** The country-wise distribution of glaucoma cases reported to the CDC VAERS.*

|  | Vaccine |  |  |
| --- | --- | --- | --- |
|  | ad26.cov2.s | BNT162b2 | mRNA-1273 |
|  | Count | Count | Count |
| Australia | 0 | 1 | 0 |
| Austria | 0 | 1 | 0 |
| Belgium | 1 | 6 | 0 |
| Czech Republic | 0 | 3 | 0 |
| Denmark | 0 | 0 | 1 |
| Estonia | 0 | 4 | 0 |
| Finland | 0 | 2 | 0 |
| Foreign (non-specific) | 0 | 1 | 2 |
| France | 1 | 18 | 1 |
| Germany | 1 | 10 | 1 |
| Great Britain | 0 | 12 | 0 |
| Greece | 0 | 2 | 0 |
| Ireland | 0 | 1 | 0 |
| Italy | 0 | 8 | 1 |
| Japan | 0 | 13 | 0 |
| Latvia | 0 | 1 | 0 |
| Lithuania | 0 | 1 | 0 |
| Netherlands | 0 | 3 | 1 |
| Norway | 0 | 0 | 2 |
| Philippines | 0 | 1 | 0 |
| Poland | 0 | 1 | 0 |
| Portugal | 0 | 2 | 0 |
| Sweden | 0 | 1 | 0 |
| Taiwan | 0 | 2 | 2 |
| Unknown | 0 | 3 | 2 |
| USA | 1 | 33 | 14 |

\* The state-wise distribution of the cases in the United States was included in the VAERS database. The origin of a report from foreign (non-US) countries was identified on the basis of unique identification number assigned by the reporting regulatory body or vaccine manufacturer.

## Acknowledgements

None

## REFERENCES

1. Mathieu E, Ritchie H, Ortiz-Ospina E, et al. A global database of COVID-19 vaccinations. Nat Hum Behav 2021;5:947–953.

2. Centers for Disease Control. The Vaccine Adverse Event Reporting System (VAERS) Request. United States Dep Heal Hum Serv (DHHS), Public Heal Serv (PHS), Centers Dis Control / Food Drug Adm (FDA), Vaccine Advers Event Report Syst 2022. Available at: https://wonder.cdc.gov/vaers.html.

3. Weinreb RN, Aung T, Medeiros FA. The Pathophysiology and Treatment of Glaucoma: A Review. JAMA 2014;311:1901. Available at: /pmc/articles/PMC4523637/ [Accessed May 18, 2022].

4. Nickells RW, Howell GR, Soto I, John SWM. Under Pressure: Cellular and Molecular Responses During Glaucoma, a Common Neurodegeneration with Axonopathy. http://dx.doi.org/101146/annurev.neuro051508135728 2012;35:p153–179. Available at: https://www.annualreviews.org/doi/abs/10.1146/annurev.neuro.051508.135728 [Accessed May 18, 2022].

5. Tham YC, Li X, Wong TY, et al. Global Prevalence of Glaucoma and Projections of Glaucoma Burden through 2040: A Systematic Review and Meta-Analysis. Ophthalmology 2014;121:2081–2090.

6. Jayaram H, Strouthidis NG, Gazzard G. The COVID-19 pandemic will redefine the future delivery of glaucoma care. Eye (Lond) 2020;34:1203–1205. Available at: https://pubmed.ncbi.nlm.nih.gov/32405050/ [Accessed June 6, 2022].

7. Anon. Welcome to MedDRA | MedDRA. Available at: https://www.meddra.org/ [Accessed May 18, 2022].

8. Centers for Disease Control and Prevention (CDC). CDC WONDER FAQs. Available at: https://wonder.cdc.gov/wonder/help/faq.html#8 [Accessed May 18, 2022].

9. Centers for Disease Control and Prevention (CDC). The Vaccine Adverse Event Reporting System (VAERS) Data Request. Available at: https://wonder.cdc.gov/controller/datarequest/D8 [Accessed June 7, 2022].

10. Fedak KM, Bernal A, Capshaw ZA, Gross S. Applying the Bradford Hill criteria in the 21st century: how data integration has changed causal inference in molecular epidemiology. Emerg Themes Epidemiol 2015;12:14. Available at: /pmc/articles/PMC4589117/ [Accessed June 6, 2022].

11. Ozonoff A, Nanishi E, Levy O. Bell’s palsy and SARS-CoV-2 vaccines. Lancet Infect Dis 2021;21:450–452. Available at: https://pubmed.ncbi.nlm.nih.gov/33639103/ [Accessed June 6, 2022].

12. Fowler N, Mendez Martinez NR, Pallares BV, Maldonado RS. Acute-onset central serous retinopathy after immunization with COVID-19 mRNA vaccine. Am J Ophthalmol case reports 2021;23. Available at: https://pubmed.ncbi.nlm.nih.gov/34151047/ [Accessed June 6, 2022].

13. Santovito LS, Pinna G. Acute reduction of visual acuity and visual field after Pfizer-BioNTech COVID-19 vaccine 2nd dose: a case report. Inflamm Res 2021;70:931–933. Available at: https://pubmed.ncbi.nlm.nih.gov/34086060/ [Accessed June 6, 2022].

14. Wasser LM, Roditi E, Zadok D, et al. Keratoplasty Rejection After the BNT162b2 messenger RNA Vaccine. Cornea 2021;40:1070–1072. Available at: https://pubmed.ncbi.nlm.nih.gov/34029238/ [Accessed June 6, 2022].

15. Phylactou M, Li JPO, Larkin DFP. Characteristics of endothelial corneal transplant rejection following immunisation with SARS-CoV-2 messenger RNA vaccine. Br J Ophthalmol 2021;105:893–896. Available at: https://pubmed.ncbi.nlm.nih.gov/33910885/ [Accessed June 6, 2022].

16. Rallis KI, Ting DSJ, Said DG, Dua HS. Corneal graft rejection following COVID-19 vaccine. Eye (Lond) 2022;36. Available at: https://pubmed.ncbi.nlm.nih.gov/34426655/ [Accessed June 6, 2022].

17. Ivanov K, Garanina E, Rizvanov A, Khaiboullina S. Inflammasomes as targets for adjuvants. Pathogens 2020;9.

18. Bayas A, Menacher M, Christ M, et al. Bilateral superior ophthalmic vein thrombosis, ischaemic stroke, and immune thrombocytopenia after ChAdOx1 nCoV-19 vaccination. Lancet (London, England) 2021;397:e11. Available at: https://pubmed.ncbi.nlm.nih.gov/33864750/ [Accessed June 6, 2022].

19. Papasavvas I, Herbort CP. Reactivation of Vogt-Koyanagi-Harada disease under control for more than 6 years, following anti-SARS-CoV-2 vaccination. J Ophthalmic Inflamm Infect 2021;11. Available at: https://pubmed.ncbi.nlm.nih.gov/34224024/ [Accessed June 6, 2022].

20. Wu P, Duan F, Luo C, et al. Characteristics of Ocular Findings of Patients with Coronavirus Disease 2019 (COVID-19) in Hubei Province, China. JAMA Ophthalmol 2020;138.

21. Gupta A, Madhavan M V., Sehgal K, et al. Extrapulmonary manifestations of COVID-19. Nat Med 2020;26.

22. Li JPO, Lam DSC, Chen Y, Ting DSW. Novel Coronavirus disease 2019 (COVID-19): The importance of recognising possible early ocular manifestation and using protective eyewear. Br J Ophthalmol 2020;104.

23. Behera G, Jossy A, Deb AK, et al. Spontaneous suprachoroidal haemorrhage in haemophilia coincident with ChAdOx1 nCoV-19 vaccine. Eur J Ophthalmol 2022:112067212210982. Available at: https://pubmed-ncbi-nlm-nih-gov.ezp-prod1.hul.harvard.edu/35484818/ [Accessed June 6, 2022].

24. Wagle A, Wu B, Gopal L, Sundar G. Necrosis of uveal melanoma post-COVID-19 vaccination. Indian J Ophthalmol 2022;70:1837. Available at: https://journals.lww.com/ijo/Fulltext/2022/05000/Necrosis_of_uveal_melanoma_post_COVI D_19.87.aspx [Accessed June 6, 2022].

25. Wang MTM, Niederer RL, McGhee CNJ, Danesh-Meyer H V. COVID-19 vaccination and the eye. Am J Ophthalmol 2022;240:79–98. Available at: http://www.ajo.com/article/S0002939422000708/fulltext [Accessed June 5, 2022].

26. Anon. Health Insurance Portability and Accountability Act of 1996 (HIPAA) | CDC. Available at: https://www.cdc.gov/phlp/publications/topic/hipaa.html [Accessed May 19, 2022].

